# NHS Hospital ‘Learning from Deaths’ reports: A qualitative and quantitative analysis of the first year of a countrywide patient safety programme

**DOI:** 10.1101/2020.10.15.20213132

**Authors:** Zoe Brummell, Cecilia Vindrola-Padros, Dorit Braun, S. Ramani Moonesinghe

## Abstract

**Introduction:** Potentially preventable deaths occur worldwide within healthcare organisations. Organisational learning from incidents is essential to improve quality of care. In England, inconsistencies in how NHS secondary care trusts reviewed, investigated and shared learning from deaths, resulted in the introduction of national guidance on ‘Learning from Deaths’ (LfDs) in 2017. This guidance provides a ‘framework for identifying, reporting, investigating and learning from deaths’. Amendments to NHS Quality Account regulations, legally require NHS trusts in England to report quantitative and qualitative information relating to patient deaths annually. The programme intended trusts would share this learning and take measurable action to prevent future deaths.

**Method:** We undertook qualitative and quantitative secondary data analysis of all NHS secondary care trust LfDs reports within their 2017/18 Quality Accounts, to review how organisations are using the LfDs programme to learn from and prevent, potentially preventable deaths.

**Results:** All statutory elements of LfDs reporting were reported by 98 out of 222 (44%) trusts. The percentage of deaths judged more likely than not due to problems in healthcare was between 0% and 13%. The majority of trusts (89%) reported lessons learnt; the most common learning theme was poor communication. 106 out of 222 trusts (48%) have shared or plan to share the learning within their own organisation. The majority of trusts (86%) reported actions taken and 47% discussed or had a plan for assessment of impact. 37 out of 222 trusts (17%) mentioned involvement of bereaved families.

**Conclusions:** The wide variation in reporting demonstrates that some trusts have engaged fully with LfDs, while other trusts appear to have disengaged with the programme. This may reveal a disparity in organisational learning and patient safety culture which could result in inequity for bereaved families. Many themes identified from the LfD reports have previously been identified in national and international reports and inquiries. Further work is needed to strengthen the LfDs programme.

## INTRODUCTION

Globally, adverse events while receiving medical treatment are a leading cause of morbidity and mortality.[1] From studies within the United States and Europe the percentage of ‘preventable’ or ‘potentially preventable’ deaths is likely to lie somewhere between 0.5% and 8.4% of hospital deaths.[2-6] In England between April 2017 and end of March 2018 there were 299,000 deaths occurring in hospital or within 30 days of discharge.[7] This amounts to an estimate of between 1,495 and 25,116 potentially preventable deaths in England in 2017/2018. There is a moral imperative for healthcare organisations to learn from these deaths and take measurable action to prevent potentially preventable deaths. Healthcare organisations are made up of individuals who have the ability to learn: however organisational learning is ‘more than the sum of individual learning’ and is distinct from unreflective action taking.[8] It is more than simply creating change for change’s sake, as an ‘illusion of learning’.[9] Organisational learning is the ability to apply knowledge and understanding to increase effective organisational action.[8, 10] Effective organisational learning is crucial to improve patient safety and probably requires both safety-I (understanding why things go wrong) and safety-II (understanding why things go right) approaches.[11, 12] In addition central regulation and performance management may have some effect on improving care, but quality improvement, leadership, public engagement, proper resourcing, education, and training are needed for a safer health service.[12]

In April 2016 an independent review demonstrated a lack of systematic approach and meaningful change in response to unexpected deaths at Southern Health NHS Foundation Trust.[13] The Care Quality Commission (CQC), which is responsible for monitoring, inspection and regulation of healthcare services within England, conducted a wider review into the investigations of deaths. They found inconsistencies in the way NHS trusts became aware of, investigated and shared learning from deaths.[14] In response, the NHS launched a new programme of work to improve standards. This included national guidance on Learning from Deaths’ (LfDs), providing a ‘framework for NHS Trusts on identifying, reporting, investigating and learning from deaths in care’. The objectives of the guidance included supporting the NHS in England to develop an understanding of why deaths contributed to by problems in care happen, with the aim of ensuring that findings are shared and acted upon, to prevent recurrence.[15] In July 2017 guidance was published on implementing the LfDs framework at trust board level,[16] and amendments to statutory regulations followed. These changes made annual reporting of both quantitative and qualitative information relating to patient deaths a legal requirement in England.[17] The reporting mechanism was built into the NHS “Quality Accounts” system – where NHS secondary healthcare providers are legally required to produce a publicly available annual report about the quality of their services.[18] Guidance was not given on expected number of deaths, how to judge if a death was more likely than not due to problems in care, or on examples of learning, actions or how to assess impact of any actions. It was instead left to individual NHS trusts to decide how they would undertake and process these requirements. Guidance was given that NHS trust board leadership should ‘share relevant learning across the organisation and with other services where the insight gained could be useful’.[15] and that trusts should ‘engage meaningfully with bereaved families and carers’.[19] It was not a statutory requirement to report on bereaved family and carer engagement or to report sharing of learning. Given the lack of consistency that led to establishing the LfDs programme, this study sets out to analyse LfDs reporting, to ascertain if trusts are reporting as legally required, to evaluate the quality of reporting, and to understand if organisational learning in its truest sense is occurring.

## METHODS

This is a qualitative and quantitative study of an NHS safety improvement programme. We undertook analysis of 2017/2018 quality account data from NHS secondary healthcare trusts in England. We excluded ambulance trusts (they are not required to report until 2020/2021). This study has been reported using Standards for Reporting Qualitative Research.[20]

Our objectives were to describe the quality of reporting, and to thematically analyse the reports to derive key learning for the NHS and beyond. We undertook analysis of LfDs as set out in the 2017 amendment to the NHS 2010 quality account regulations.

Our evaluation of the quality of reporting involved review of compliance of reports against regulation numbers 27.1 to 27.6 (table 1).[17] Where trusts did not fully report we sought to understand why this may have been the case from comments within the quality account itself. Data not found from the trust 2017/2018 quality account was not included in the analysis.

**Table 1:** NHS Quality Accounts LfDs Regulations [17].

| **Regulation number** | **Summary of regulatory requirement** |
| --- | --- |
| **27.1** | The number of patients who have died during the annual reporting period |
| **27.2** | The number of the deaths (in 27.1) that have undergone a case record review or investigation |
| **27.3** | An estimate of the number of deaths in 27.2 which the provider judges to be more likely than not to have been due to problems in care, with explanation of method to assess this |
| **27.4** | What the provider has learnt from reviews/investigations in relation to deaths (in 27.3) |
| **27.5** | A description of the actions the provider has taken or will take in response to what they have learnt |
| **27.6** | An assessment of the impact of the actions (from 27.5) |

In addition to statutorily required reporting we also looked for evidence within the 2017/18 LfDs report of family/carer engagement, which included evidence of involvement in learning and/or addressing family/carer concerns and/or appointing family liaison officer or similar as a result of a patient death. We also looked for evidence of sharing LfDs incidents both within the trust and more widely (for example with other organisations).

Quantitative analysis was undertaken and reported using descriptive statistics.

Qualitative evaluation to derive key learning themes was undertaken through document analysis using content and thematic analysis.[21] In order to collect data systematically, we first identified initial LfDs learning and action themes for reporting, and then developed a classification system for these. The first investigator (ZB) reviewed and analysed twenty 2017/2018 quality accounts, undertook open coding and combined this with information presented at the NHS Improvement London Network for Learning from Deaths event (October 2018), where themes (mixed learning and action) from London Trusts were discussed. Following the initial review, we reviewed the further 202 NHS trust 2017/18 quality accounts. Each account was reviewed by the same reviewer twice to ensure full data capture. The process of bracketing to reduce subjective analysis was utilised.[22] During data capture further themes emerged, were modified, merged and changed iteratively. Recurring themes were identified using a method of exploratory data analysis,[23] coding, identification of themes, recoding and using frequency charts. Data were captured in Microsoft excel.

### Patient and public involvement

This study forms part of a larger programme of work which is overseen by a public and relatives steering group to improve relevance from the perspective of those affected by deaths in healthcare and to reduce biases from the healthcare staff researchers. The steering group have been involved in the planning, design and development of conclusions, through face-to-face meetings and email correspondence. The involvement of a steering group member in authoring this paper has significantly and positively influenced the reporting of this study, ensuring focus on reporting family involvement. The authors reflect that PPI has been essential to this study to ensure that the views of bereaved family members were central to the concerns examined. The reporting of patient and public involvement (PPI) has been undertaken using guidance for reporting involvement of patients and the public 2 – short form (GRIPP2-SF).[24]

## RESULTS

Quality accounts were reviewed for all 222 secondary care trusts in England.

### Quality of Reporting

98 out of 222 (44%) trusts reported all six statutory elements of the LfD reporting framework. Two trusts did not report any parts of the LfDs regulatory requirements.[25, 26] The total number of deaths reported (regulation 27.1) varied from 3 deaths to 7756 deaths [27, 28] The number of case record reviews or investigations undertaken relative to the number of patient deaths in individual trusts varied between 0.2% and 100% of deaths; the average was 43.7%.

There was variation between 0 and 13% in the number of deaths which the provider judged to be more likely than not to have been due to problems in care. 22 trusts did not report any figure in this section of the quality accounts, reasons given for this included:

- ‘data collection challenges’[29]s
- ‘unable to provide a reliable figure’[30]
- ‘we do not carry out investigations with a view to determining whether the death was wholly or partly due to problems in the care provided’[31]
- ‘currently no research base on this for mental health services and no consistent accepted basis for calculating this data’[32]

111 out of 222 trusts (50%) noted the use of Structured Judgement Reviews (SJRs) (either Royal College of Physicians or Royal College of Psychiatrists) either alone or in combination with other forms of investigation or review to assess problems in care.[33] Trusts not using SJRs used a variety of other methods including: Confidential Enquiry into Stillbirths and Deaths in Infancy (CESDI) framework, Root Cause Analysis (RCA) and PReventable Incidents Survival and Mortality (PRISM) methodology.[34, 35]

Regulation 27.4 asks trusts to describe ‘what the provider has learnt from reviews/investigations in relation to deaths’ where this was related to deaths which the provider judged to be more likely than not to have been due to problems in care (regulation 27.3). 25 out of 222 trusts (11%) did not report any lessons learnt from deaths; of these 25 trusts, 9 trusts had reported 1 or more death judged to be more likely than not due to problems in care, the other 16 trusts had either reported zero deaths judged to be more likely than not due to problems in healthcare or had not reported. However, 49 out of 222 trusts (22%) which reported that they had no deaths judged more likely than not due to problems in care, also reported lessons learnt, many caveating this with an explanation that they had learnt valuable lessons through the process of case note review/investigation.

Trusts were asked to undertake ‘a description of the actions the provider has taken or will take in response to what they have learnt’ (Regulation 27.5). 30 out of the 222 trusts (14%) did not report any actions taken as a result of learning. One trust reported that they felt they were ‘at too early a stage of development to be able to take actions from specific learning’.[36]

Regulation 27.6 asked trusts to undertake ‘an assessment of the impact of the actions’. 105 out of 222 trusts (47%) discussed assessment of impact. This includes trusts that had a plan of any sort including a future plan. Several trusts used audits and/or quality improvement projects to check that actions are implemented. One trust stated ‘Many of these actions are difficult to objectively assess in terms of their impact as they may relate to rare occurrences, which are difficult to meaningfully audit’.[37] The 47% of trusts who had a plan for assessment of impact does not include trusts that acknowledge the need to assess the impact but stated that it was too early to be able to undertake this (or words to this effect).[36, 38] Some trusts have reported the results of the assessment of impact that they have already undertaken.[39] Several trusts appear to have misunderstood, for example reiterating the purpose of the LfDs programme, instead of assessing impact.[40, 41]

### Evidence involvement of family/carers in learning

In the 2017/18 LfDs reports 37 out of 222 trusts (17%) mentioned the involvement of families/carers either in the investigation process or in shared learning or that they communicate with/support/engage/consider families/carers after a patient dies.[42-44] A good example of working with families from one trust LfDs report states: ‘The Trust continues to learn the importance of communication with families after a death has occurred and that through meaningful engagement after a death by inviting them to contribute to the terms of reference for investigations a more detailed, meaningful and richer account of the person’s care and treatment is realised’.[45] One trust LfDs report discusses that as an action undertaken they sought to gain better education and training for staff about the importance of positive family engagement through expert external training.[46] 38 Trusts (17%) discussed as an ‘action’ that they plan to work with/communicate with/engage/support families/carers. Many of these trusts are the same trusts already undertaking family/carer engagement.

### Evidence learning shared more widely

In the 2017/18 trust LfDs reports 106 out of 222 trusts (48%) have shared or plan to share the learning more widely within their own organisation, through a variety of communication mediums: Face to face meetings or events, trust intranet (as case studies, safety alerts, newsletters).[36, 44, 47] 17 out of 222 (8%) trusts have shared or plan to share the learning outside their organisation, with neighbouring trusts or other national organisations.[47-50]

### Key Findings from the Reports Lessons learnt

The most common learning themes from all trusts who reported learning can be found in table 2. An overview of the themes arising can be found in the frequency table (figure 1).

**Table 2.** The 5 most common learning themes across all trusts.

| **Learning Themes** | **% of trusts citing theme** |
| --- | --- |
| Poor communication (including language barrier and problems with handover) | 46% |
| Problem in recognition and escalation of deteriorating patients | 42% |
| End of life planning or treatment escalation planning not evident/incomplete | 42% |
| Problems with documentation | 41% |
| Lack of/problem with risk assessment/interventions | 25% |

**Figure 1.**
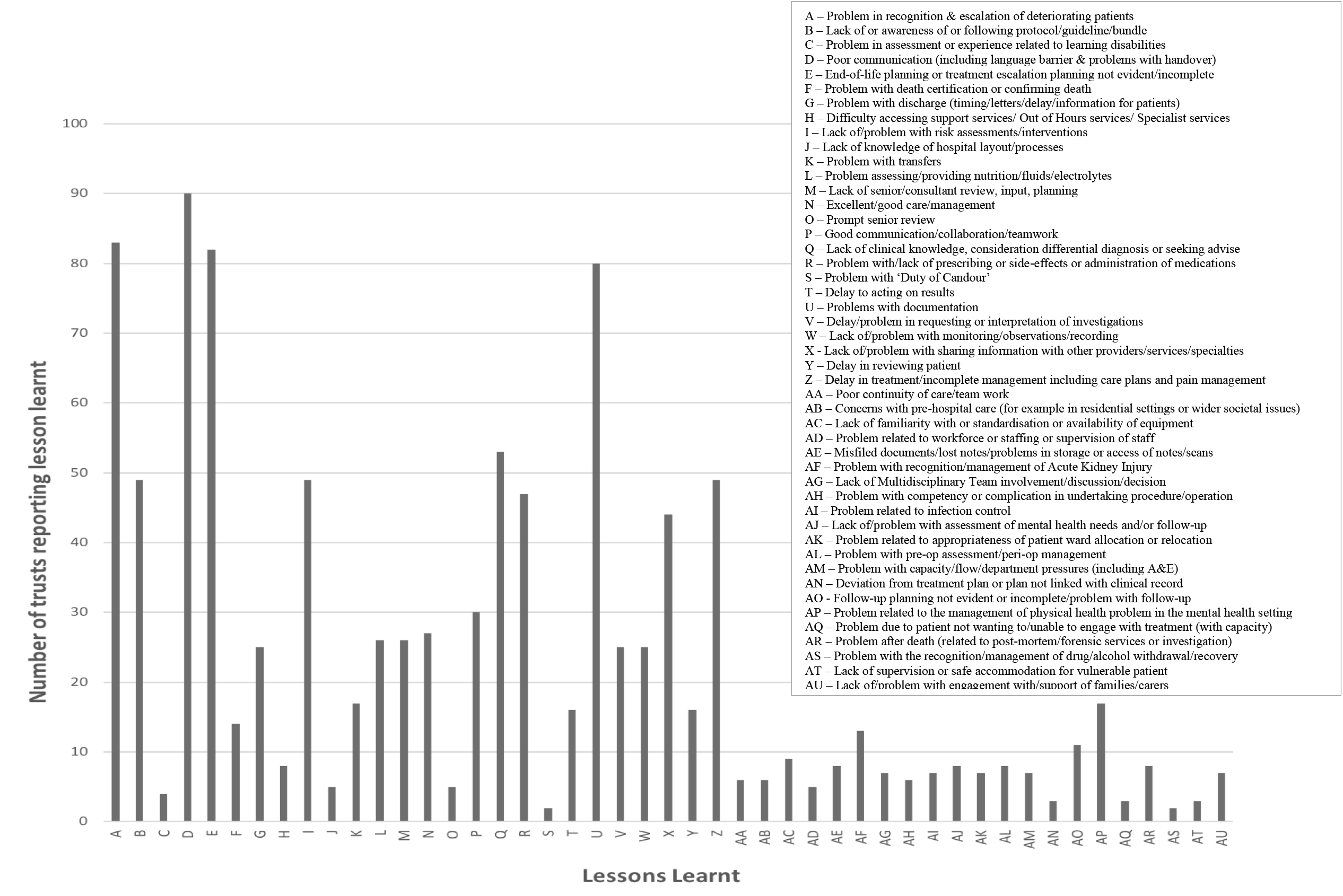
Frequency table of lessons learnt (all trusts; n=222)

Some trusts have undertaken analysis of their learning and described common themes.[51] Some have gone into great detail.[52] Others have described a specific case or cases.[53] Some trusts have identified learning and actions together, without differentiating the learning from the action. The lack of structure in reporting makes it difficult to always understand exactly what the problem was leading to the learning. This could reduce the transferability of the learning.[54] Some trusts identified ‘Good practice’ as learning points.[55] Occasionally trusts did not necessarily learn from patient deaths, but from the overall LfDs process.[56]

### Actions taken or planned to be taken

The most common action themes from all trusts who reported actions can be found in table 3. An overview of the themes arising can be found in the frequency table (figure 2).

**Table 3.** The 5 most common action themes across all trusts.

| **Action Themes** | **% of trusts citing theme** |
| --- | --- |
| Review of process/Standard Operating Procedure (SOP)/pathway | 58% |
| Highlight or produce guidelines/protocols/policies/treatment bundles/toolkits | 43% |
| Implementation programme of training/education | 43% |
| Quality improvement work or similar | 39% |
| Work to improve communication/collaboration/shared learning | 28% |

**Figure 2.**
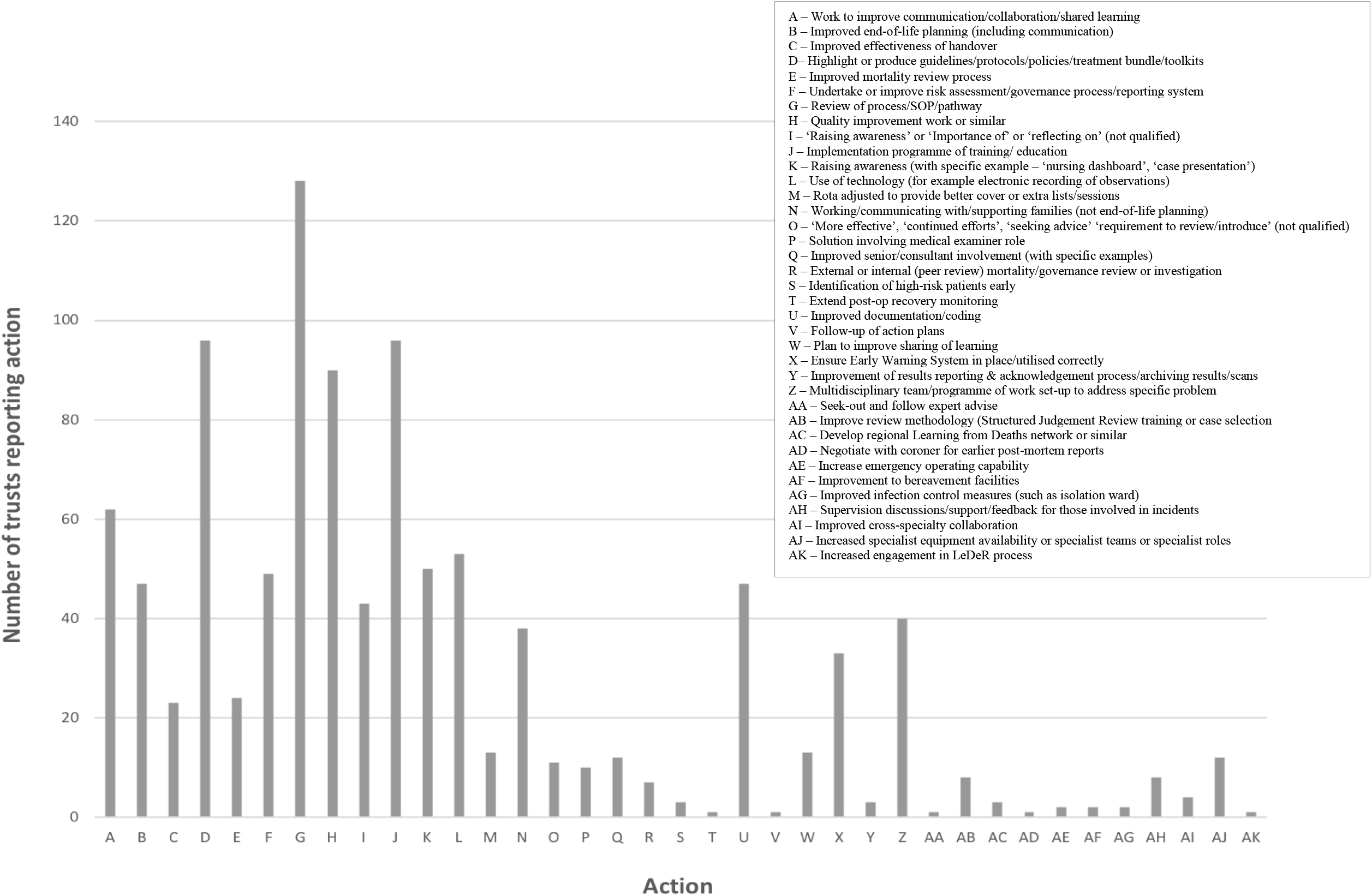
Frequency table of actions taken (all trusts; n=222)

The level of detail with regards to actions taken varies greatly with some trusts listing some specific actions as bullet points.[39] Others trusts have described a specific case or cases.[57, 58]

## DISCUSSION

This study demonstrates wide variation in both the quality of reporting and the findings from LfDs reports. Considering this is a new programme, introduced part-way through 2017/18, with limited guidance, the overall findings are somewhat encouraging. Nearly all trusts reported at least one or more element of the LfD reporting framework. Most trusts reported lessons learnt and/or actions taken, while less than half discussed assessment of impact. The lessons learnt were varied. The most common learning theme was poor communication, with the most common action being; review of process/SOP/pathway.

### Quality of Reporting

Reporting variation may be due to differences in interpretation of the guidance. There is no direct financial penalty for a trust not reporting some or all elements of the LfDs statutory requirements in their Quality Accounts. A penalty arises during CQC inspections, through assessment of implementation of LfDs.[59, 60]

The different approaches taken by trusts and the heterogeneity of data makes comparison difficult. The variation in the percentage of deaths being reviewed/investigated may be due to some trusts not having the capacity to review/investigate cases, collect and/or report accurately. Trusts with a very small number of deaths may find it easier to review all deaths than very large trusts. Some trusts have had mortality review processes in place for several years and have already been reviewing/investigating deaths, making implementation of the LfDs process easier since the structure for reviewing cases and personnel required are already in place. Some trusts may have felt at risk from negative attention by declaring total numbers of deaths and deaths judged more likely than not due to problems in care. Many trusts did however report despite the same risk. It is clear from the LfDs reports that several trusts, particularly some mental health and community trusts, did not feel that the guidance applied to them, however other similar trusts were able to comply with reporting. The results could suggest guidance was written with acute trusts in mind and perhaps need to be reconsidered for non-acute trusts. Similar findings were noted by the CQC in their report ‘Learning from deaths: A review of the first year of NHS trusts implementing the national guidance’.[60]

The variation in deaths judged more likely than not due to problems in care is larger than those noted in previous studies.[2-6] It seems unlikely than many trusts would experience no deaths judged more likely than not due to problems in care. This could realistically be the case in specialist trusts where the absolute number of total deaths is very small, or community trusts with no inpatient beds, but seems unlikely in large acute trusts. Despite the improbability several acute trusts did report zero deaths judged more likely than not due to problems in care. Further work to understand why these trusts reported zero deaths should be undertaken.

The element of the statutory LfDs reporting that prompted poor responses from most trusts was ‘An assessment of the impact of the actions’ and describing how they would undertake this. The vast majority of trusts have answered this in a vague manner. Improvements could be made by issuing further specific guidance in relation to this element of the reporting. Of the trusts who did manage to implement actions and assess impact this was often using quality improvement measurements. The use of quality improvement methodology is felt to be an important overall indicator of quality by the CQC.[61] Guidance on evaluating the impact of interventions is widely available.[62, 63]

Collectively within the LfDs reports, there is much learning, some appearing to result in impactful actions and high-level organisational learning.[8] This learning could potentially be usefully shared across the NHS and internationally. Some NHS trusts appear to have disengaged with the programme, with incomplete or partial LfDs reporting. This study suggests a lack of shared learning from the LfDs reports particularly across organisations and a lack of family engagement, despite NHS guidance.[19] Since the involvement of families and sharing learning were not statutory requirements of LfDs reporting, they may be underrepresented in the LfDs reports, this should be investigated further. The apparent disparity in organisational learning and safety culture, results in inequity for families/carers. This should be addressed by the DHSC and associated national bodies. Since the oversight bodies which were established to support the programme in its initial stages have now been stood down this seems unlikely to happen.[64]

### Key Findings from the Reports

Overall consistency with regards to identifying, reporting, investigating, learning from deaths in care and taking action has improved across most trusts. Many trusts have effectively described lessons learnt and actions taken. The continual process of learning, action and reflection which characterises effective organisational learning is essential to ensure safer healthcare and a safety culture.[65, 66] Evidence of effective organisational learning from trust LfDs reports is limited. Only a small number of trusts did not report any learning, suggesting that most trusts were able to engage with this aspect of reporting. Most of the LfDs report recommendations or actions are fairly non-specific; further detail of actions and their measurable impact would be helpful.

It is of concern that many of the lessons and recommended actions from LfDs reports have previously been identified in national and international reports and inquiries, looking at the problems associated with preventable deaths. Similar problems found in this study are also highlighted in these reports; poor clinical monitoring, poor recognition of the deteriorating patient, diagnostic errors, poor communication, lack of end of life planning, lack of information sharing between services, inadequate drug and fluid management.[67-75] This suggests many of the same problems reoccur and that healthcare systems do not effectively learn from previous failings and adds weight to the proposition that the NHS as a whole cannot become a learning organisation.[76] In view of this, it is reasonable to question whether the learning arising from LfDs reporting will result in meaningful change. If LfDs findings and recommendations are not implemented, systemic redundancy in the initiative is implied. While individual healthcare practitioners do need to take some responsibility, trusts and the DHSC should look at systems, such as institutional accountability and LfDs programme oversight to optimise outcomes. This lack of change adds to the growing body of evidence suggesting that traditional approaches to organisational learning in healthcare, such as learning from when things go wrong (safety-I) have limited effect and may suggest a role for increased learning from the patients who survive against the odds (safety-II).[77]

### Recommendations

In view of the findings from this study, in order to improve reporting quality, our recommendations are as follows:

- A more structured LfDs reporting template, including all regulatory requirements should be implemented through the quality accounts
- NHSE/I specific guidance should be developed on how trusts can undertake ‘an assessment of the impact of the actions’
- To reinstate LfDs robust regulatory reporting oversight in addition to CQC inspections

In order to improve ‘learning and action’ from deaths, our recommendations are:

- Annual collection and collation of all trust LfDs reporting for wider sharing
- Further investigation into how trusts currently involve bereaved families and carers
- Investment in leadership and support for NHS staff to enable a safety culture

### Study Limitations

This is an analysis of the first year of LfDs reporting, many trusts may not have got completely to grips with it yet. On first review the data for 2018/19 does appear to be more robust.

It is important to understand that trusts may be undertaking elements that were not statutory reporting requirements and raises the question of whether public reporting is reflective of trust engagement with the LfDs programme.

## CONCLUSION

This research shows that the LfDs programme has improved the way that NHS trusts identify, report, investigate and learn from deaths in care. However, more could be done to enhance and strengthen the programme impact, and to assess whether LfDs reporting reflects trust LfDs engagement and organisational learning.

On the basis of findings from the 2017/18 LfDs reports, national programmes led by multidisciplinary healthcare practitioners should be developed to tackle the most common problems which may have contributed to patient deaths. In the first instance programmes tackling the following issues should be developed or strengthened:

- Improving communication
- Involvement of families in care and in learning
- Processes to share learning (locally and nationally)

Further work is needed to understand which actions taken by trusts result in the biggest impact and for this learning to be shared. While LfDs can be difficult and emotive it is fundamental that healthcare systems ensure effective learning and impactful change occur.

## Data Availability

Data for this study is publicly available from the 2017/18 NHS Quality Accounts

## Footnotes

### Contributors

ZB conceived and designed the study, undertook analysis, and drafted the manuscript. All authors provided critical input to the design, analysis, interpretation of the data and revised the manuscript critically. The corresponding author attests that all listed authors meet authorship criteria and that no others meeting the criteria have been omitted. ZB is the guarantor.

### Ethical approval

This study is an analysis of publicly available data and does not require ethical approval.

### Data sharing

Data for this study is publicly available from the 2017/18 NHS Quality Accounts

### Transparency statement

We confirm that the manuscript is an honest, accurate, and transparent account of the study being reported; that no important aspects of the study have been omitted; and that there are no discrepancies from the study as originally planned.

### Dissemination to participants and related patient and public communities

We plan to disseminate results to the medical director, director for Patient safety and national medical examiner at NHS England/Improvement, the medical director at the Care Quality Commission and to the families involved in the NHS England LfDs steering group.

## Acknowledgements

We would like to acknowledge the work of the members of the Learning from Deaths: Learning and Action (LfDLaA) Public and Relatives steering group in this research.

